# Attitudes Towards Digital Health Technology: Introducing the Digital Health Scale

**DOI:** 10.1101/2021.09.03.21262482

**Authors:** Ridley Cassidy

## Abstract

**Objective:** The study sought to investigate the relationship between attitude towards digital health technology and age, gender and frequency of use of digital health technology and to consider whether age, gender and frequency of use present potential barriers to accessing future healthcare in the UK. Differences in technological affinity are likely to lead to differences in the adoption of digital health technology and subsequent inequalities in healthcare between older and younger people and between men and women.

**Design:** The study represents an example of a technology adoption study employing a survey-based cross sectional correlational design. Attitude towards digital health technology was measured using the 20 item Digital Health Scale. Age, gender, frequency of use of health technology and employment status data were gathered using a demographics questionnaire. The opportunity sample (N = 247) included volunteer participants aged 16-84 years (*M* = 31.7, *SD* = 19.35, 156 females and 91 males).

**Results:** Results indicated a significant negative correlation between age and positive attitude towards digital health technology (*r* = -0.24, *p* < .01). Gender differences in attitudes towards digital health technology were non-significant (*p* > .05). Significant differences in frequency of use were also found, where occasional and frequent use resulted in more positive attitudes than never having used digital health technology (*p* < 0.05) and participants reporting frequent use were significantly older than those reporting never or occasional use (*p* < .05)

**Conclusion:** Findings identified age, but not gender, as a significant factor in attitude towards digital health technology, suggesting that continued and increased reliance on digital technology in healthcare may lead to age, but not gender, related inequalities in access to healthcare in the UK. That frequent users of digital health technology were also older, highlights the greater demand for healthcare services by older individuals and is further evidence for the potential of digital healthcare to lead to age related inequalities in access to and provision of healthcare. Recommendations for successful application of digital healthcare technology are considered in the light of these findings.

## Introduction

Technology is ever present in all aspects of our lives and will be for as long as it continues to develop new ways to help us live more efficiently and improve our quality of life. Technology’s ongoing growth and development at an irrepressible rate has “transformed the way we live, work and communicate” (Maitland, 2012). Therefore we must embrace the growth of technology and advance with it in all areas of our lives.

Health technology is defined as “the application of organized knowledge and skills in the form of devices, medicines, vaccines, procedures and systems developed to solve a health problem and improve quality of lives” (World Health Organization, 2017).The majority of technological advances so far in healthcare are mainly due to new opportunities created by the Internet that challenge traditional health care (Eysenbach, 2001), the impact of which continues to grow. Technology and its continued development offer great potential for increased efficiency for communication, even more so with advances in mobile communication and accessibility of mobile devices. A study conducted by The Office of Communications (Ofcom, 2016) showed that 93% of adults in the United Kingdom owned a mobile phone, 71% of those owned a smartphone, offering huge potential for the healthcare sector to exploit, and in doing so, revolutionise the way healthcare is delivered.

### Emerging Health Technologies

AXA PPP Healthcare have published a recent report detailing a number of examples of work exploring the potential of digital healthcare technology (AXA PPP Healthcare, 2017). Examples include a patient-focused digital services study where SMS (short message service) reminders were employed as part of a smoking cessation programme, with results producing a “9% 52-week quit rate compared to 4% with the over-the-counter nicotine replacement therapy and 1% quit rate with simple GP advice”. The AXA PPP Healthcare report goes on to refer to a study directed by the Hurley Group, London, which demonstrated the potential that digital health services offered. A series of patient orientated online services, including specialist advice, symptom checking, self-help content and GP consulting, resulted in half of those patients who used the ‘eConsult’ service being managed remotely, shorter waiting times and fewer GP appointments. To explore the potential of these services on a larger scale and establish its future role in the healthcare system, the service has now been extended to one million patients. Patient-focused digital services are now adopted nationally by health service providers as an effective method of communicating with and monitoring patients (AXA PPP Healthcare, 2017).

Digital health takes several forms. eHealth^1^ is one of the most rapidly developing and adopted forms of digital health technology. Whilst it has no single unifying definition, perhaps because of its endless possibilities, Eyesenbach (2001) defines it as “technical development, but also a state-of-mind, a way of thinking, an attitude, and a commitment for networked, global thinking, to improve health care locally, regionally, and worldwide by using information and communication technology”. The benefits of eHealth are noted by Eysenbach and include increased efficiency in health care and better quality of care. The presence of eHealth continues to grow globally within the healthcare sector, its benefits are becoming widely acknowledged, “The findings suggest that Americans’ believe that health IT adoption is an effective means to improve the quality and safety of health care.” (Gaylin, Moiduddin, Mohamoud, Lundeen, & Kelly, 2011). Chiron Health also refer to the proven value of telemedicine for individuals in assisted living facilities, where it allows doctors to manage cases by remote visits and determine if a hospitalisation is actually necessary (Chiron Health, 2017).

Telemedicine^2^ is an example of a fast emerging digital health technology. It involves the remote diagnosis and treatment of patients by means of telecommunications technology which can “speed up diagnoses, increase access to care for remote populations, reduce health care costs and even relieve physician shortages.” (Parks, 2017). Telemedicine is still in the early stages of development but has already been adopted worldwide by a number of health organisations to deliver efficient health care and exploit the associated economic benefits. For example, in America the cost of a telemedicine appointment is around “$45 compared with approximately $100 for the same appointment in a doctor’s office” (Beck, 2016). Recent statistics illustrate how this method of health care has already been widely adopted in America, with more than 15 million Americans receiving some kind of medical care remotely in 2015, a figure that the American Telemedicine Association expect to grow by 30% in 2017 (Beck, 2016).

Particularly pertinent in the context of the ongoing COVID 19 global pandemic is that telemedicine also “allows for better care in places where medical expertise is hard to come by” and reduces the patient’s exposure to potentially contagious patients, reducing the risk of the spread of diseases within the hospital walls (Beck, 2016). Parks (2017) reports the findings of two survey studies that help demonstrate the extent to which technology is being used by health professionals to deliver healthcare services, the first, an MGMA Stat poll exploring if physicians will or currently offer telehealth services in 2017, found that almost 17 percent of the 1,325 respondents said that they already offer telemedicine services and a further 21 percent have plans to use the technology.”, and the second, telehealth Index survey from American Well, found that 57 percent of physicians are interested in seeing patients over video if clinically appropriate. Together these results provide evidence for an irreversible shift in the way we deliver healthcare as the worldwide adoption of telemedicine and similar emerging digital health technologies becomes ever more apparent.

Other digital health technologies include 3D bioprinting^3^, microchips modelling clinical trials^4^, optogenetics^5^ and digestible sensors^6^, as well as many others still in the very early stages of development. All of these digital health technologies “are starting to allow healthcare practitioners to offer cheaper, faster and more efficient patient care than ever before” (Honigman, 2014).

### Barriers to Digital Health Technology

The e adoption of digital health technologies for the delivery of health care does present some potential disadvantages, including a breakdown in the relationship between health professional and patient, a breakdown in the relationship between health professionals, and issues concerning the quality of health information (Hjelm, 2005). Such issues may present significant obstacles for the adoption of digital health as the standard for health care delivery.

Equally, people’s attitude has been highlighted as another factor with the potential to slow down or prevent the introduction of digital health (Arning & Ziefle, 2009). According to the ABC Model, attitudes are defined as “relatively enduring organization of beliefs, feelings, and behavioral tendencies towards socially significant objects, groups, events or symbols” (Hogg & Vaughan 2005, p. 150) that have an **A**ffective component (feelings and emotions), a **B**ehavioural component (how a person’s attitude influences how they think they will behave) and a **C**ognitive component (knowledge and beliefs). The ‘principle of consistency’ states that there is an expectation that a person’s attitude and how they behave are consistent with each other (McLeod, 2014). A study conducted by *HealthMine* to investigate telemedicine awareness, reported 39% of participants were not aware of telemedicine, with participants who had never used telemedicine, 42% preferred in-person doctor visits (Beck, 2016). These findings illustrate how attitudes and awareness offer valuable insight into behavior, highlighting a barrier to the adoption of health technology, not just for users but also for providers. A survey containing 1500 family physicians established that only 15% of the practices had adopted telemedicine as part of their long term future strategy (Beck, 2016), signifying that telemedicine has a long way to go before becoming a universally accepted and adopted health technology and highlighting the need for further research in the area of digital health technology adoption.

Despite the well documented benefits of new technology, not everyone is able to experience these benefits equally. The ‘digital divide’^7^ in the use of general technology has been reported in areas such as education (Digital Divide in UK Schools, 2017) and business (Anderson, 2016) and limits the impact of technology because of such things as geographical location, socioeconomic factors, and age and gender disparities (Digital Divide in the UK, 2017). It is reasonable to suggest that such factors may also affect digital health technology, limiting its adoption and resulting in inequalities in healthcare. There are relatively few technology adoption studies focussing specifically on digital health technology in the UK, but the ones there are suggest that, amongst a number of socioeconomic factors, age and gender may be major factors in the adoption of health technologies. Arning and Ziefle (2009) explored the role played by age and gender in the acceptance of health technology and found that older participants perceived higher ‘utilization barriers’ and reported higher concerns about data safety issues than younger participants. A 2015 Health Tech Nation Survey, conducted by AXA PPP Healthcare, confirmed increasing adoption of health technology but identified clear generational differences between attitudes to, and the adoption of, different types of health technology (AXA PPP Healthcare, 2017). Younger people seem keener to embrace ‘big data’ and demonstrate a belief that health technology can help with the management of long-term conditions (especially obesity, diabetes, cancer and heart disease), while those over 55 have more of a focus and interest in health monitoring devices.

Kaushal, Ancker, Silver and Miller (2013) reported a relationship between age and attitudes towards health technologies, suggesting age as a potential barrier to digital health, “Belief that EHR [Electronic Health Records] would improve healthcare quality was significantly more common among those under the age of 40 years than other age groups.” Similar trends were found in the Care Innovations study which suggesting that “People over 65 are traditionally less-experienced with technology when compared to younger adults, and most people over 65 show declines in perceptual, motor, and/or cognitive function that can interfere with their ability to use technologies.” (Care Innovations, 2013).

A recent study exploring acceptance of eHealth technologies in older people living with chronic pain, reported that older adults wanted eHealth to be delivered alongside existing in-person visits from health and social care professionals (Currie, Philip & Roberts, 2015), The results suggest that although older participants are open to the idea of the change in the way health care is delivered, they do not want their health to be entirely dependent on health technology. These results highlight age as a potential barrier to the adoption of digital health technology, supporting the need for further research in this area.

The idea of gender as a possible barrier in the adoption of technology has been apparent for some time, since gender disparities were reported in general technology acceptance studies (e.g. Bowser & Daugherty, 1998). In a study of public health promotion, Ek (2013) reported that “findings show clearly that being female is a strong predictor of being more proactive and engaged in seeking, gaining and discussing health-related issues”. Ek concluded that to succeed in public health promotion and interventions, the measures taken should be much more sensitive to the gender gap in health information behaviour. This identifies gender as a potentially important factor in understanding the use of digital health technology. However, there is contradictory evidence regarding gender differences, with some studies reporting inconclusive relationships between gender and attitudes towards health technologies. Kaushal et al. (2013), for example, found “none of the sociodemographic variables [investigated] were significantly associated with privacy and security concerns about HIE [Health Information Exchange].” In light of inconclusive findings from existing studies, gender differences in attitudes towards health technology will be further investigated in the present study.

As general technology studies have reported frequency of use as a significant correlate of attitudes towards technology (*r* = .49, *p* < 0.0005, Lukow, 2005), it will also be considered in the present study as a factor related to potential barriers to the adoption of digital health technology. In a study exploring public attitudes towards digital health technology, Gaylin, Moiduddin, Mohamoud, Lundeen and Kelly (2011) found that affinity to health technology was affected by how frequently individuals used health technology, “respondents who are more experienced with electronic technology have more favourable attitudes toward health IT [information technology].” A further study investigating patient attitudes towards mobile phone-based health monitoring revealed that those who were more comfortable with mobile phone based monitoring (mHealth) owned and used a smart phone prior to the study, suggesting an association between use of technology and attitudes to and adoption of digital health (McGillicuddy, et al., 2013).

### The Present Study

Potential barriers limiting access to healthcare have serious implications and present a potential public health risk. Thus, it is important to examine the role that age and gender play in attitude towards digital health technology and, in turn, in the adoption of digital healthcare technology. This is particularly important in light of the increasing aged population in the UK, “70% of UK population growth between 2014 and 2039 will be in the over 60 age group, an increase from 14.9 to 21.9 million people” (Government Office for Science, 2016), and associated higher demands of older people on healthcare services, “Most people aged 75 and over have one or more health conditions” (Mortimer & Green, 2015). Consequently, there is an increasing need for older people to use digital health technology to access healthcare services. If older people are unwilling or unable to engage with digital health technology, this creates and inequality in healthcare that could have a major negative impact on the health of the population in the UK.

The present study examines the relationship between age, gender and the attitudes towards digital health technology. To the author’s knowledge there is no suitable existing measure of attitudes to digital health. Using a newly developed attitude towards digital health technology scale (The Digital Health Scale, DHS) focussing on knowledge of, emotional response to, and behaviour towards health technology (i.e., the ABC model of attitudes, McLeod, 2014), the study explores age and gender as possible barriers to access to digital health and, in the broader context, access to healthcare. The study tests the hypotheses that there will be a significant correlation between age and attitude towards digital health technology and that there will be a significant difference between males and females in their attitude towards health technology. Reported frequency of use is also expected to reveal differences in attitude towards digital health technology and age. Investigating these factors, the study addresses the broader issue of whether digital health technology is likely to improve access to healthcare consistently across the population. The aim is to identify factors that will help eradicate or lessen the risk of inequalities in health care and ensure equality in delivery of health care in the UK.

## Method

### Design

The study used a survey-based cross sectional correlational design. The study variables were age in years, gender, frequency of us of health technology (never, occasionally, frequently), occupation (student, employed, retired, other) and attitudes towards digital health technology measured using the self-report Digital Health Scale (digitalhealthscale.com). The following controls were employed to counter the effects of extraneous variables^8^: participants under 16 years were excluded from the study as they are not responsible for their own healthcare; the survey was anonymous to reduce the effects of social desirability, a form of response bias^9^, and increase the validity of participants’ responses; the survey includes both positively and negatively worded statements and reverse scoring of some statements to limit the effects of acquiesce bias^10^.

### Participants

The study adopted a non-probability convenience sample of 247 volunteers recruited from a college population of faculty, staff and students to participate in the study. The sample included 91 males and 156 females with an age range of 16 to 84 years (*M* = 31.7, *SD* = 19.35). Occupational status was 131 students, 77 employed, 26 retired, 13 other. Frequency of use of health technology was 101 never, 119 occasionally, 27 frequently.

### Materials

#### The Digital Health Scale (DHS)

The DHS is a self-report measure of attitudes towards digital health technology (digitalhealthscale.com). It contains 20 statements relating to how people feel, behave and think about health technology. Statements represent the different components of attitudes, i.e., emotion, behaviour and cognition (McLeod, 2014) and the different aspects and issues that are relevant to the use of technology in health, including ease of use, reliability and privacy. Participants respond to each statement along a 5-point Likert scale (McLeod, 2008) from strongly agree to strongly disagree depending how much they agree or disagree with the statement. Example statements include ‘I don’t really understand how to use heath technology’ and ‘If I am ill, I prefer to look on the internet for help than go to the doctors’. A total DHS score is achieved by summing the scores of the 20 individual statements, giving a theoretical scoring range of 20-100. There are a mix of positively and negatively worded statements, with scores for positive statements (2,4,5,9,11,13,14,16,18) reversed so that a high DHS score represents a positive attitude towards digital health.

The DHS was piloted with a focus group of five members of the general public who agreed to take part (age range 16-48 years; 3 females, 2 males). Members completed a 23 item draft version of the survey and provided verbal feedback as part of a group discussion. Following piloting the wording of three statements was revised, three items were deleted, and a definition of digital health technology was added in order to make it statements more understandable and relatable for the participants, ‘*the use of electronic systems and devices, including smartphones and apps, computers, telephone booking systems and the internet, for diagnosis and treatment of illnesses and more general healthcare organisation and education’*.

The remaining 20 statements constituted the final Digital Health Scale used in the study. The internal reliability of the final 20 item DHS was assessed using Cronbach’s item-analysis. The alpha coefficient was .84, which shows that the reliability of the scale is acceptable (Field, 2013).

#### Demographics Questionnaire

A short questionnaire will be used to collect age, gender, frequency of use of digital health technology and employment status data from participants.

#### Procedure

The study was approved by Winstanley College Institutional Review Board. Following college ethics approval, a request for study volunteers together with the participant information sheet and survey link were distributed via email to staff and students at a college in the United Kingdom. Participants were fully informed of the purpose of the study and that participation was voluntary anonymous. They were advised that by completing and returning the survey they were giving their informed consent to participate in the study. To make recruitment and data collection more efficient the DHS and demographic questions were available online using the online survey platform SurveyMonkey©. There was the option to complete a print version of the survey if participants preferred. Volunteers who were willing to participate completed the online or print survey and demographic questions. Online responses to were captured automatically in an Excel spreadsheet. Participants completing print copies returned these anonymously to a collection point.

## Results

A Shapiro-Wilk’s Test (*p* > .05), as well as visual inspection of the histogram, showed that attitudes towards digital health technology response data were approximately normally distributed, with skewness of -0.283 (*SE* = 0.155) and a kurtosis of 0.167 (*SE* = 0.309). Therefore, parametric data analysis techniques can legitimately be used for most parts of the analysis involving DHS scores.

### Digital Health and Age

Figure 1 suggests a moderate negative correlation between attitude towards digital health and age. Pearson’s correlation coefficient confirmed a significant small to moderate (Cohen, 1988) correlation (*r*(247)= -.32, 2-tailed, *p* < .01). Therefore the experimental hypothesis is accepted and the null hypothesis is rejected. There is a significant negative correlation between age attitudes towards health technology.

**Figure 1.**
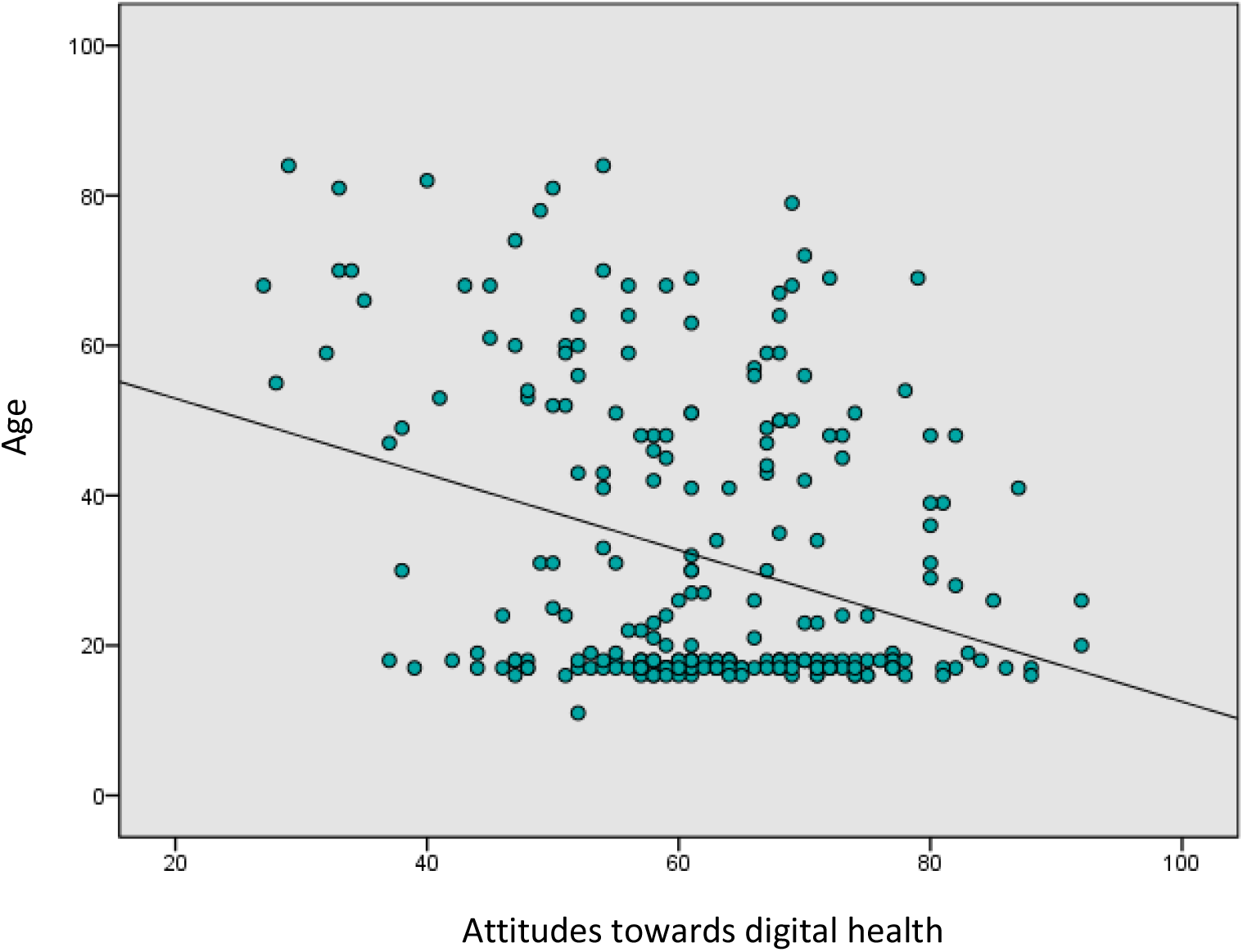
Attitudes towards digital health by age

As age data failed to approximate normal distribution (Shapiro-Wilk’s test *p* < 0.05, skewness = 1.05, *SE* = 0.155; kurtosis = -0.227, *SE* = 0.309), a non-parametric Spearman’s correlation was also calculated. Although the correlation coefficient was slightly smaller, a significant negative correlation was nevertheless confirmed. (*r*(247)= -.24, 2-tailed, *p* < .01).

### Digital Health and Gender

Figure 2 shows a small difference in mean attitudes towards digital health scores between males (*M* = 62.8, *SD* = 12.6, *N* = 91) and females *(M* = 61.6, *SD* = 12.2, *N* = 156). Levene’s test for equality of variance was non-significant (*p* > .05), therefore homogeneity of variance can be assumed. An independent samples t-test conducted to compare the effect of gender on attitudes towards digital health confirmed the difference was non-significant (*t*(245) = 0.467, 2-tailed, *p* > .05).

**Figure 2.**
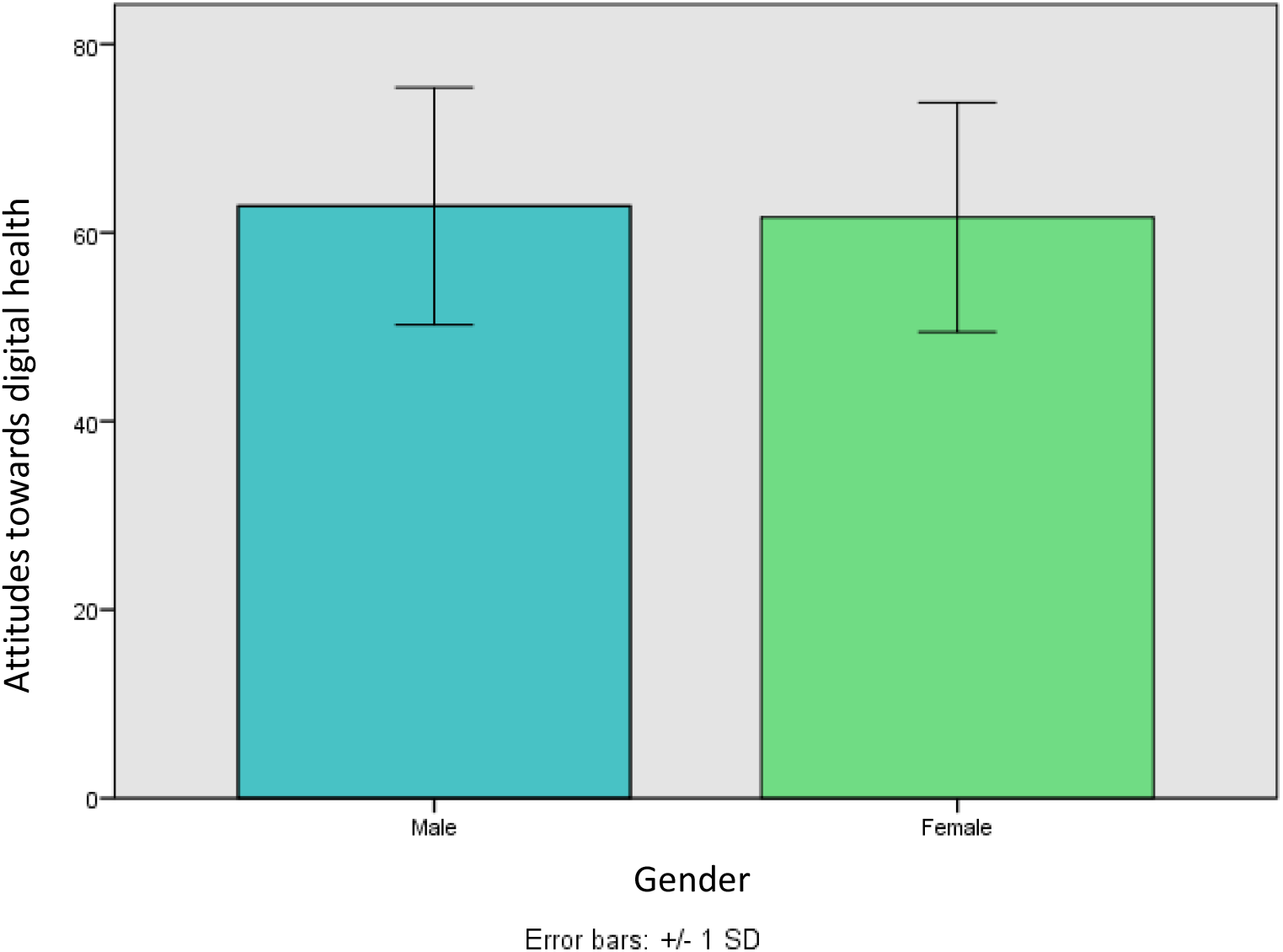
Attitudes towards digital health by gender

### Health Technology and Use

Figure 3 shows a difference in mean attitudes towards digital health scores between the three self-reported levels of use of digital health, *never* (*M* = 59.1, *SD* = 12.1, *N* = 101), *occasionally* (*M* = 64.1, *SD* = 11.6, *N =* 119), *frequently* (*M* = 64.3, *SD* = 14.4, *N* = 27). A one-way between subjects analysis of variance (ANOVA) conducted to compare the effects of frequency of use on attitude towards digital health in never, occasionally and frequently conditions was significant (*F*(2, 244) = 5.056, *p* < .01). Post hoc comparisons using the LSD test confirmed a significant difference in mean scores between never and occasionally (*p* < 0.01) and between never and frequently (*p* < 0.05) conditions. The difference between occasionally and frequently conditions was non-significant (*p* > .05). Thus, there is a significant difference in attitude towards digital health according to frequency of use of digital health.

**Figure 3.**
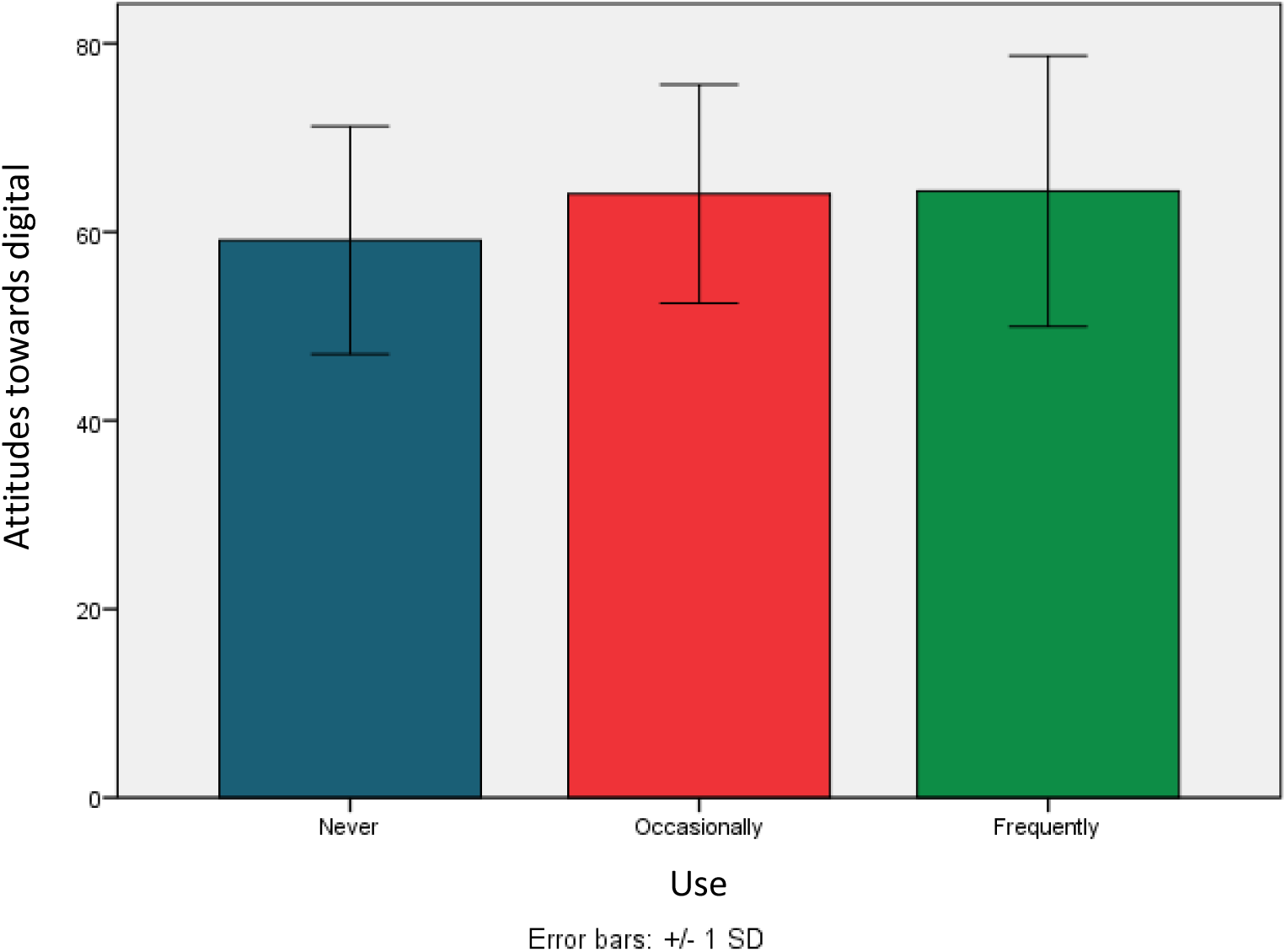
Attitudes towards digital health by frequency of use

Figure 4 shows a difference in mean age according to the level of self-reported frequency of use of digital health, *never* (*M* = 26.3, *SD* = 18.3, *N* = 101), *occasionally* (*M* = 34.5, *SD* = 19.2, *N =* 119) and *frequently* (*M* = 39.4, *SD* = 19.5, *N* = 27). A one-way between subjects ANOVA conducted to compare the effects of frequency of use on age in never, occasionally and frequently conditions was significant (*F*(2, 244) = 7.618, *p* < 0.01). Post hoc comparisons using the LSD test confirmed a significant difference in mean age between never and occasionally (*p* < 0.05), never and frequently (*p* < 0.05), and between occasionally and frequently (*p* < 0.05) conditions. Thus, there are significant differences in mean age according of frequency of use of digital health technology.

**Figure 4.**
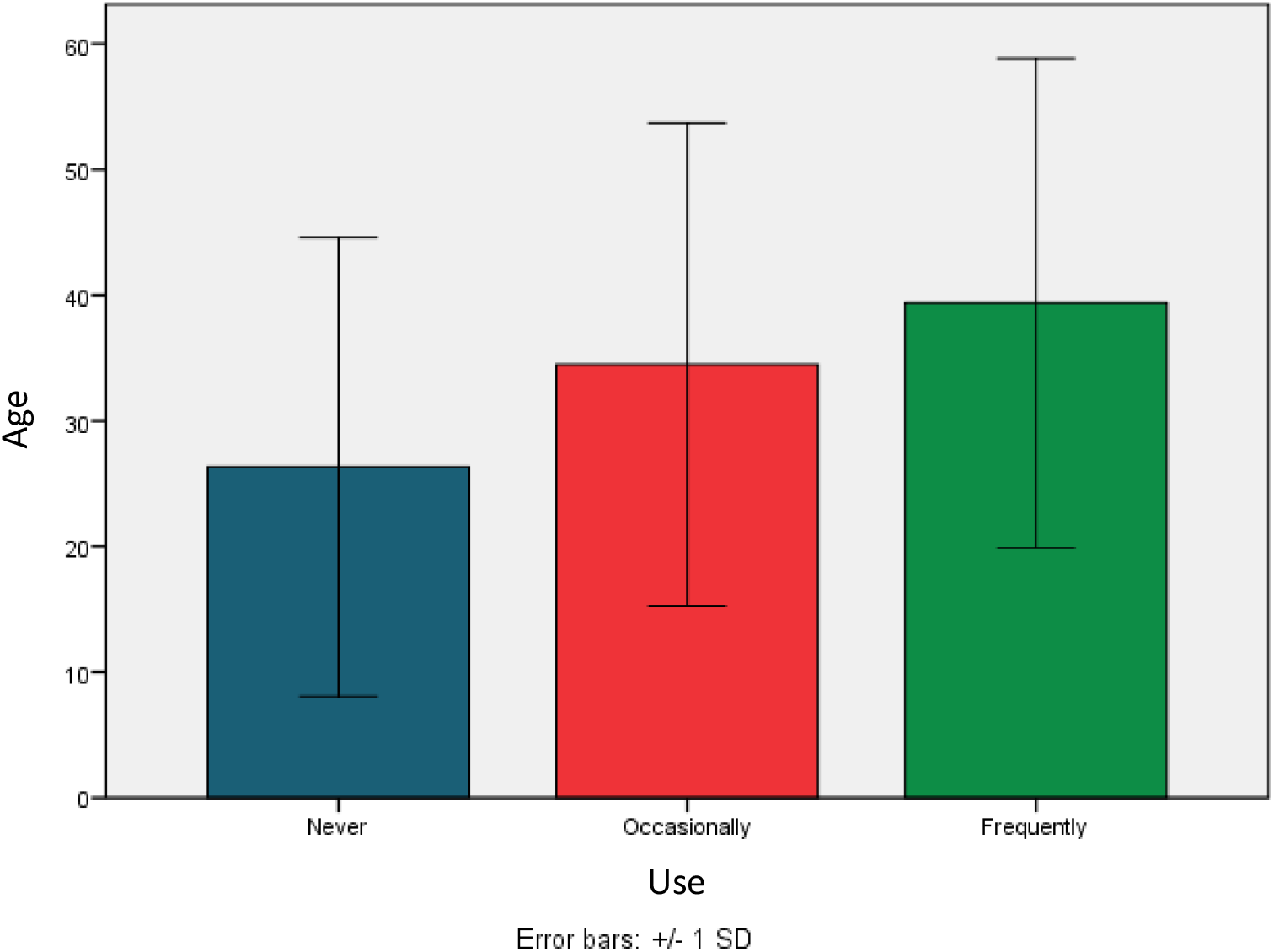
Mean age by frequency of use of digital health

### Digital Health and Occupation

Figure 5 shows a difference in mean attitude towards digital health between occupation types, *student* (*M* = 64.0, *SD* = 10.6, *N* = 131), *employed* (*M* = 63.2, *SD* = 12.7, *N* = 77), *retired* (*M* = 50.8, *SD* = 13.7, *N* = 26) and *other* (*M* = 57.9, *SD* = 11.9, *N* = 13). A one-way between subjects (ANOVA) conducted to compare the effects of occupation type on attitude towards digital health was significant (*F*(244, 2) = 7.618, *p* < 0.01). Post hoc comparisons using the LSD test confirmed a significant difference in mean attitude towards digital health between retired and student (*p* < .001) and between retired employed (*p* < 0.001). No other comparisons were significant (*p* > .05). Thus, there are significant differences in age according to occupation.

**Figure 5.**
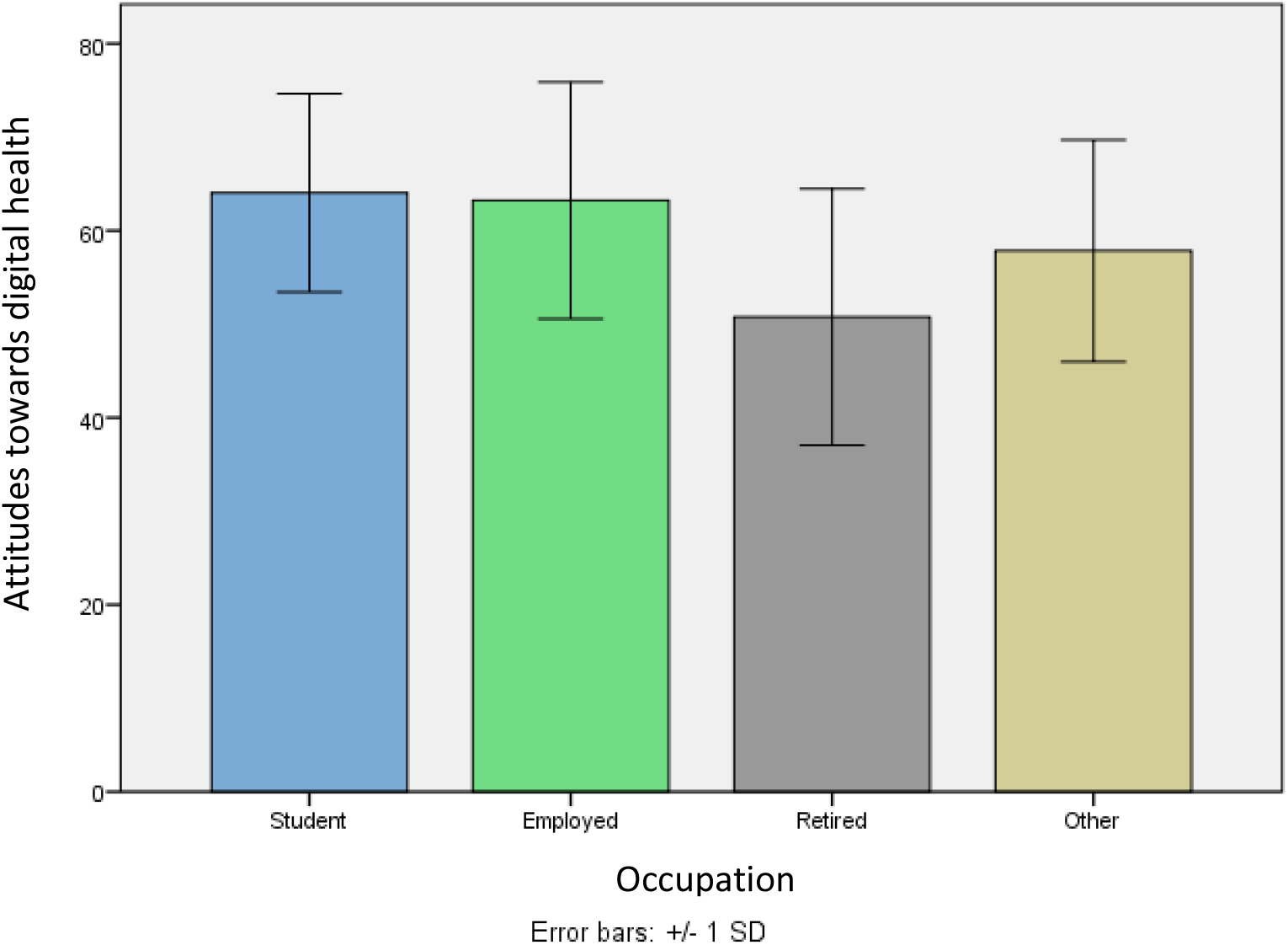
Attitudes towards digital health by occupation

## Discussion

The aim of the study was to explore the relationship between age, gender and attitude towards digital health, having identified age and gender as potential barriers to the adoption of digital health and therefore limiting access by some people in the UK to healthcare services. Because of the rapidly ageing population in the UK (Government Office for Science, 2016) and the greatly increased needs and demand on healthcare services of older people (Mortimer & Green, 2015), there was a particular focus in the study on age and the adoption of digital health. The link between attitudes and behaviour is well-established, where behaviour is consistent with attitude (McLeod, 2014), and there is evidence highlighting attitudes as an important factor in technology adoption studies (e.g., AXA PPP Healthcare, 2017; Kaushal, Ancker, Silver, & Miller, 2013). Therefore, in the absence of a suitable attitudinal measure, the study developed the Digital Health Scale (digitalhealthscale.com) as a measure of attitudes towards digital health to investigate age and gender differences in the likely acceptance and adoption of digital health in the UK.

### Digital Health Technology and Age

Based on previous research that identifies age as a factor affecting attitudes towards, and adoption of, technology in general (Arning & Ziefle, 2009), it was predicted that there would be a significant relationship between age and attitudes towards digital health. Findings from the present study show a significant small to moderate negative correlation between age and attitude towards digital health. Older people therefore expressed a more negative attitude towards digital health. When occupation data was explored, retired people expressed a significantly more negative attitude towards digital health than both students and employed people. Assuming that retired people are generally older—the average age of retirement in the UK for males is 65 years and 64 years for females (Organisation for Economic Co-operation and Development, 2017)—this further supports the association between age and attitude towards digital health. Age and use of digital health were also explored to investigate which age group used digital health most frequently. Findings showed that the group reporting using digital health frequently were significantly older than those reporting using it occasionally or never using it, supporting the suggestion that older people have a greater demand for healthcare services (Mortimer & Green, 2015) so are increasingly having to use digital health as the main, or only, way to access services. These findings are in line with a number of existing general technology and health technology studies. For example, a German study exploring the acceptance of technology in relation to age, concluded that older participants tended to feel “intimidated by these new health-technologies, as they cannot appraise the consequences of using these devices for their living context” (Arning & Ziefle, 2009), suggesting how older people feel uncomfortable when using technology. A 2017 report, The Impact of health technology on people’s behaviours (AXA PPP Healthcare, 2017), also included findings that demonstrated how younger people have a more positive attitude towards digital health, valuing it more and are more willing to engage with it, than older people. Other studies, including Kaushal et al. (2013) and Currie and Roberts (2015), exploring age as a potential barrier to the adoption of digital heath reported similar findings, concluding that age does play a role in attitudes towards and acceptance of digital health.

### Digital Health and Gender

While findings are inconsistent, some previous technology adoption studies have reported gender differences. Therefore, gender was assessed in the present study for its impact on attitudes towards digital health. Findings did not support a significant gender difference in attitude towards digital health.

The findings were not completely unexpected given that existing research investigating gender differences in technology studies is inconclusive. Findings from recent studies that explored gender disparity in relation to technology are divided. Some studies report that gender does have an impact on attitudes towards both general technology health technology, suggesting how females have a more negative attitudes in some areas of technology use (Bowser & Daugherty, 1998) but are more interested in seeking health related information (Ek, 2013). Similar trends were not evident in the data of the present study, perhaps partly due to the difference in the aims and design of the study, but the absence of gender differences is also reported in other technology adoption studies, including Kaushal, et al (2013), which helps support findings from the present study.

### Use of Digital Health

Because the primary focus of the study was to explore adoption of digital health and because the rationale for the study rested heavily on the suggestion that older people have a greater need to access health services and therefore a greater need to use digital health, self-reported usage data was collected. Findings showed that those participants reporting frequent use of digital health were significantly older than those who reported never or occasional use. This finding provides support for the notion presented in the study and supported in published evidence (McGillicuddy, et al., 2013; Gaylin et al., 2011) that older people, because of their declining health and need for health care, currently have a greater need to use digital health as a means to access health care services.

The growing aged population (Government Office for Science, 2016; Green, 2015) and the seemingly irreversible trend towards digital health (Parks, 2017) means that the frequency of use of digital technology is likely to grow. This adds extra emphasis to the need to ensure that older people, in particular, develop an increasingly positive attitude towards digital health. Analysis of reported usage data in the present study revealed that those reporting frequent and occasional use of digital health reported a significantly more positive attitude towards digital health than those who never use digital health. This finding is supported by other studies that report usage as a significant factor in attitudes towards and adoption of technology, including Gaylin et al. (2011) and McGillicuddy, et al. (2013), who both reported similar findings suggesting that individuals who use digital health technology more, and have more experience with digital health technology, demonstrate a more positive attitude towards digital health when compared to those who use it on a less frequent basis. Similar trends are reported in the present data

Thus, it seems providing opportunities for individuals, particularly older people and those likely to avoid using digital health, is likely to help develop a more positive attitude towards digital health and lead to increased adoption of digital health. This has already been demonstrated in general technology studies, where technology educations programmes had a positive effect on attitudes towards technology (Bowser & Daugherty, 1998).

### Limitations and Directions for Future Research

The study had some limitations that should be considered when interpreting the findings. Younger participants were overrepresented in the study sample so the size of the reported correlation coefficient between age and attitudes towards health technology may have been affected by attenuation bias^11^ caused by the skew of the age data (Kendall & Stuart, 1958, cited in Barrett, 2001). So, the degree of association may be greater than reported. Data were collected using both online and face-to-face methods. The study was conducted in the UK with UK residents. This may limit the application of findings to the other populations, and conclusions can only safely be drawn for the UK. Finally, the limitations of the Digital Health Scale should be considered. Although the DHS showed acceptable internal reliability (Cronbach’s alpha .84, Field, 2013) and the reported correlation with age and significant differences between usage levels and between occupation types support the validity of the DHS as a measure of attitude towards health technology, this is a newly developed measure and further evidence of its reliability and validity is needed.

As well as further work developing and testing the DHS as a measure of attitudes towards digital health, future studies should focus on gathering empirical data on the relationship between attitudes towards digital health and actual usage. If significant associations continue to be reported then work needs to be done on how to foster positive attitudes towards digital health and if providing opportunities for ‘at risk’ groups, including older people, to use health technology in supported situations is a valuable way to improve attitudes and adoption of digital health. Studies incorporating the opinions of healthcare professionals and the users of digital health should also be undertaken.

## Conclusion

The study provides evidence supporting age, but not gender, as a significant factor explaining attitudes towards digital health. Older individuals expressed a more negative attitude towards health technology. There is a strong association between attitude and behaviour, as demonstrated in a number of technology adoption studies (e.g. Kaushal et al., 2013). Thus, the suggestion that age may be a barrier to access to healthcare, leading to a potential public health risk around inequality in health care, is worth further consideration. This is particularly important given the aging population in the UK who are likely to have a greater demand for health service provision (Government Office for Science, 2016; Green, 2015). Older individuals may be unable to, or unwilling to, access these healthcare services if, as seems to be the growing and irreversible trend (Parks, 2017), they are delivered via technology-based platforms. The study did provided evidence that older, retired people reported more frequent use of digital health technology than younger, student and employed people. This suggests that healthcare services already rely heavily on technology and the greater demand for these services by older people. Therefore, it seems critical that measures are taken to ensure older people in particular are likely to and able to access digital health services. Failing to take steps to ensure this could limit access to health care and with long-term health implications. The study also provided evidence that people who reported more frequent use of digital health technology also expressed a more positive attitude towards digital health than those who reported never having used digital health technology. This is a common finding in technology adoption studies exploring general technology, where increased use of technology and technology education programmes altered attitudes towards technology (Bowser & Daugherty, 1998; Lukow, 2005). The experience does however have to be positive, and it is therefore argued here that positive experience with digital health technology, provided as part of an organised and structured intervention targeting older people, is one way in which negative attitudes could be addressed, helping to ensure that age presents less of a barrier to access to health care. Finally, in developing the Digital Health Scale, the study provides an import and unique contribution to measurement that can be used to gain critical insight in digital health research and intervention studies focussing on user behaviour.

## Data Availability

The data that supports the findings of this study are available on request from the corresponding author.

## Acknowledgements

Thank you to Judith Wallace for support provided during completion of the project.

## Appendix The Digital Health Scale

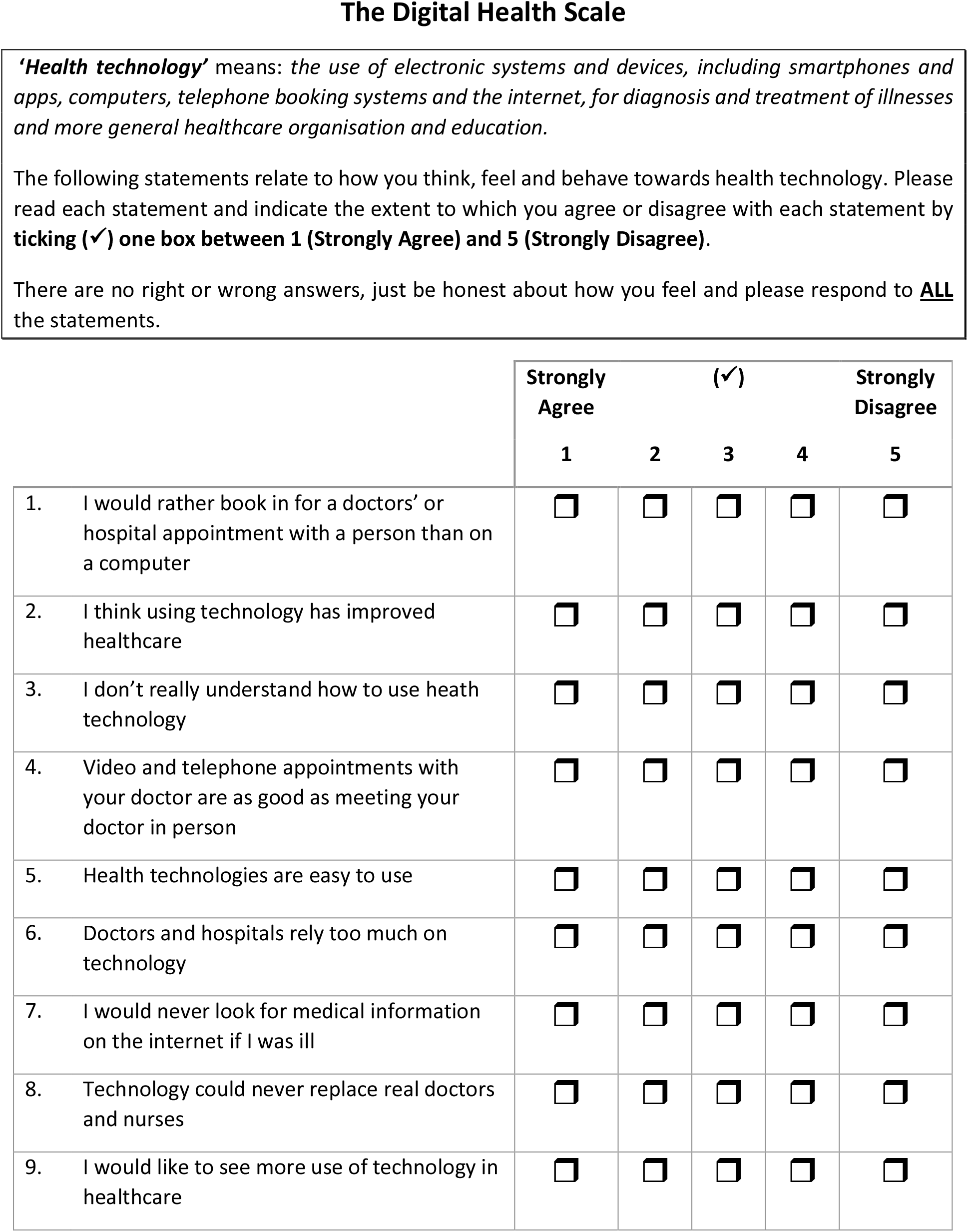
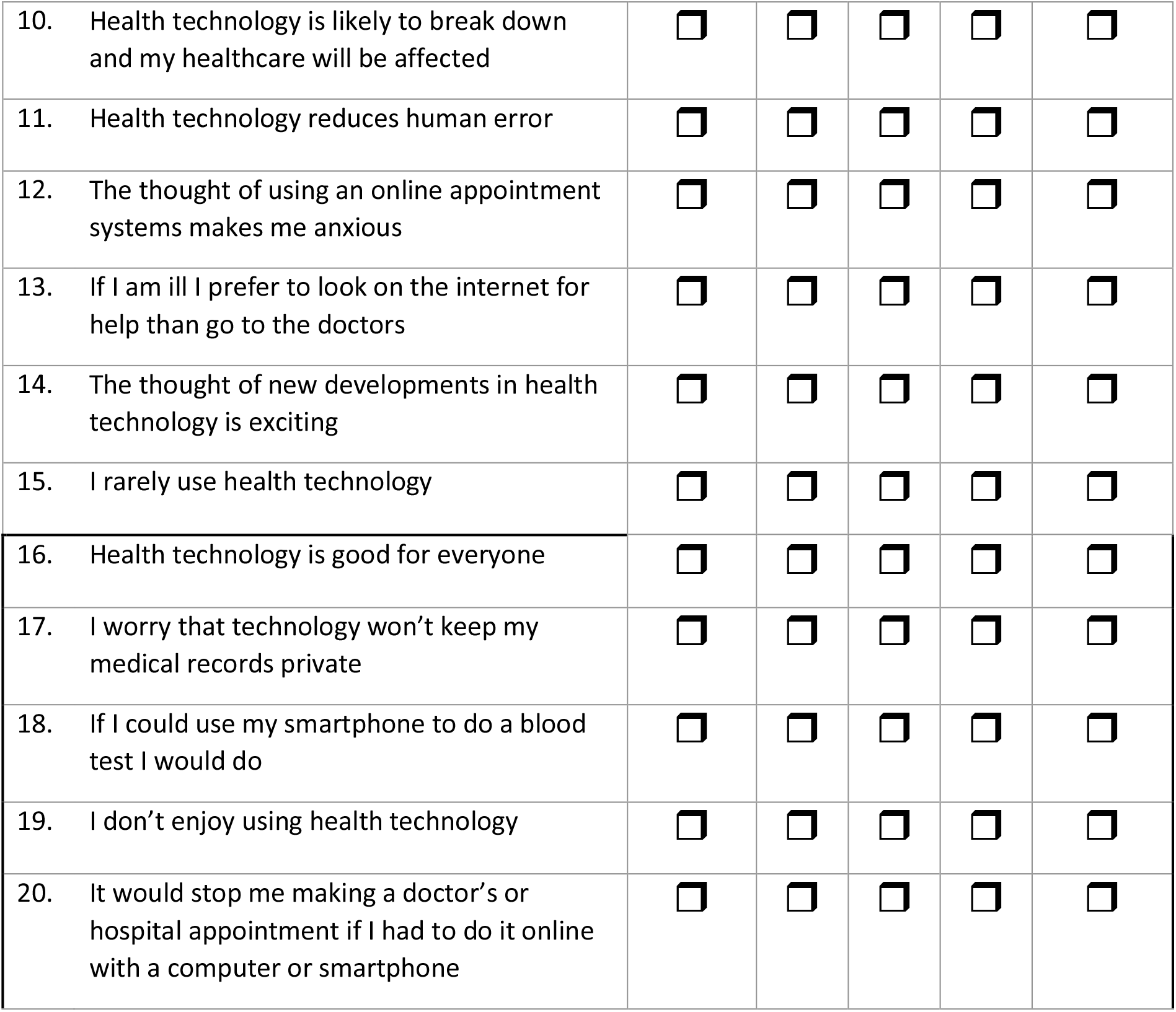

|  | Strongly Agree |  | (✓) |  | Strongly Disagree |
| --- | --- | --- | --- | --- | --- |
|  | 1 | 2 | 3 | 4 | 5 |
| 1. I would rather book in for a doctors’ or hospital appointment with a person than on a computer | ☐ | ☐ | ☐ | ☐ | ☐ |
| 2. I think using technology has improved healthcare | ☐ | ☐ | ☐ | ☐ | ☐ |
| 3. I don’t really understand how to use health technology | ☐ | ☐ | ☐ | ☐ | ☐ |
| 4. Video and telephone appointments with your doctor are as good as meeting your doctor in person | ☐ | ☐ | ☐ | ☐ | ☐ |
| 5. Health technologies are easy to use | ☐ | ☐ | ☐ | ☐ | ☐ |
| 6. Doctors and hospitals rely too much on technology | ☐ | ☐ | ☐ | ☐ | ☐ |
| 7. I would never look for medical information on the internet if I was ill | ☐ | ☐ | ☐ | ☐ | ☐ |
| 8. Technology could never replace real doctors and nurses | ☐ | ☐ | ☐ | ☐ | ☐ |
| 9. I would like to see more use of technology in healthcare | ☐ | ☐ | ☐ | ☐ | ☐ |

|  |  |  |  |  |  |  |
| --- | --- | --- | --- | --- | --- | --- |
| 10. | Health technology is likely to break down and my healthcare will be affected | ☐ | ☐ | ☐ | ☐ | ☐ |
| 11. | Health technology reduces human error | ☐ | ☐ | ☐ | ☐ | ☐ |
| 12. | The thought of using an online appointment systems makes me anxious | ☐ | ☐ | ☐ | ☐ | ☐ |
| 13. | If I am ill I prefer to look on the internet for help than go to the doctors | ☐ | ☐ | ☐ | ☐ | ☐ |
| 14. | The thought of new developments in health technology is exciting | ☐ | ☐ | ☐ | ☐ | ☐ |
| 15. | I rarely use health technology | ☐ | ☐ | ☐ | ☐ | ☐ |
| 16. | Health technology is good for everyone | ☐ | ☐ | ☐ | ☐ | ☐ |
| 17. | I worry that technology won't keep my medical records private | ☐ | ☐ | ☐ | ☐ | ☐ |
| 18. | If I could use my smartphone to do a blood test I would do | ☐ | ☐ | ☐ | ☐ | ☐ |
| 19. | I don't enjoy using health technology | ☐ | ☐ | ☐ | ☐ | ☐ |
| 20. | It would stop me making a doctor's or hospital appointment if I had to do it online with a computer or smartphone | ☐ | ☐ | ☐ | ☐ | ☐ |

## Footnotes

1 A relatively recent term for healthcare practice supported by electronic processes and communication.

2 “Telemedicine allows health care professionals to evaluate, diagnose and treat patients at a distance using telecommunications technology.” (Chiron Health, 2017).

3 Using living cells to 3D print organs (Gilpin, 2014).

4 Microchip modelling clinical trials aim to replace the use of animals in clinical trials to more accurately test the safety and efficacy of treatment for human patients and spare the lives of countless animals typically used in testing. (Honigman, 2014).

5 Neuroscientists can target a single neuron in the brain of a mouse merely by turning on a light. (Honigman, 2014).

6 A sensor that transmits information about a patient to medical professionals to help them customise the care to the individual as well as the care provided to other individuals experiencing similar health conditions or ailments. (Honigman, 2014).

7 “The idea of the “digital divide” refers to the growing gap between the underprivileged members of society, especially the poor, rural, elderly, and handicapped portion of the population who do not have access to computers or the internet; and the wealthy, middle-class, and young Americans living in urban and suburban areas who have access.” (Digital Divide, 2017)

8 Extraneous Variables are undesirable variables that influence the relationship between the variables that an experimenter is examining. (Psychology World, n.d.)

9 The bias shown by people to present themselves in positive ways to the community. (Psychology Dictionary).

10 The bias shown by people who agree with positively worded statements

11 The underestimation of the size of correlation coefficient, resulting from errors in the independent variable.

